# Sickle hemoglobinopathy research in Zimbabwe and Zambia (SHAZ): Protocol for setting up an international Sickle Cell Disease registry

**DOI:** 10.1101/2024.05.28.24308028

**Authors:** Patience Kuona, Gwendoline Q Kandawasvika, Catherine Chunda-Liyoka, Ian M Ruredzo, Pauline M Sambo, Pamela Gorejena-Chidawanyika, Hamakwa M Mantina, Takudzwa J Mtisi, Cynthia Phiri, Lawson Chikara, Natasha M Kaweme, Exavior Chivige, Jombo Namushi, Tendai Maborekeke, Uma H Uthale, Collen Masimirembwa

## Abstract

Of the 500 000 children born with sickle cell disease annually, most cases occur in Africa, contributing to significant morbidity and mortality associated with limited sickle cell disease (SCD) health outcomes data and reduced access to therapeutic plus preventive care. We aim to develop and manage a standardized electronic SCD registry, establish consistent standards of care (SoC) for patients, improve the SCD research and biobanking capacity in Zimbabwe and Zambia. This five-year program employs mixed methods that include infrastructure and skilled manpower capacity building of SCD clinics, registry, biobanking, cohort and implementation science research studies to improve SCD treatment outcomes. We are collaborating with the SickleInAfrica consortium (Ghana, Mali, Nigeria, Tanzania, Uganda, and South Africa), the African Institute of Biomedical Sciences and Technology (AiBST) and St Jude’s Children Research Hospital. We established the SCD registry in Zimbabwe and Zambia for children and adult patients enrolling 1796/4000 (45%) participants to date. We are participating in SickleInAfrica consortium research activities, training health workers and educating SCD patient communities on SoC. This collaboration with African researchers, policymakers, health workers, and SCD patient communities will improve uptake of SCD SoC and increase our research capacity.

## Introduction and background

Sickle Cell Disease (SCD) is a heterogeneous multi-organ disorder characterized by the presence of an abnormal oxygen carrier, hemoglobin S (HbS)[1]. Globally, the distribution of the sickle cell gene mirrors the geographical distribution of malaria[2]. Population migration has resulted in the spread of SCD beyond tropical regions including Southern Africa which was historically thought to have a low prevalence[3]. An estimated 500,000 children are born with the condition in the world every year with 80% of these births occurring in Africa[4]. SCD significantly contributes to under-five mortality in Africa and globally[5]. It is estimated that 50-90% of children with SCD in Africa die before their 5th birthday[6]. Very few children in Africa survive beyond 18 years of age and those who survive into adulthood have premature mortality due to end organ damage [7] e. Despite the high burden of SCD in Africa, there is limited data on the prevalence and incidence of the disease and its health outcomes. There are limited appropriate and context-specific clinical guidelines and policies for the care of affected individuals in Africa. There is also inadequate uptake of evidence-based preventive and therapeutic practices that have reduced morbidity and mortality in high-income countries[8,9].

Historically, HbS was considered to be of low prevalence in Southern Africa but the prevalence of SCD has increased over the years due to migration[3]. Zimbabwe is estimated to have received close to one million migrants from countries with a high prevalence of SCD and with approximately 100,000 being HbS carriers[3]. A 2010 modeling study estimated that the frequency of the HbS allele in Zimbabwe was 0.021 compared to 0.171 in Nigeria, 0.112 in Zambia, 0.074 in Tanzania, and 0.003 in South Africa[10]. Another modeling study in 2010 estimated that 534 babies were born with SCD in Zimbabwe with the majority dying undiagnosed early in life[11]. An investigation of beta-globin gene haplotypes in bio-banked samples of three cohorts from Malawi, South Africa, and Zimbabwe demonstrated that 12% of 50 healthy unrelated participants from Zimbabwe were carriers of the Hb S gene[12].

In Zambia, SCD is most common in the northern region bordering the Democratic Republic of Congo (DRC) where the sickle cell trait (SCT) rate was reported to be 17.5%[13,14]. A more recent study by Mkushi and Serenje reported a prevalence of 15.5% of SCT and 3.4% of SCD [15]. The precise prevalence of SCD in Zambia remains unknown due to limited epidemiological surveys that have been conducted. In 2017, SCD patients accounted for 12% of the all-cause hospital admissions at University Teaching Hospital (UTH) in Zambia[16]. SCD was ranked fourth of the top five causes of mortality and was amongst the top 10 reasons for seeking medical attention at the University Teaching Hospitals - Children’s Hospital (UTHs - CH) in Lusaka the capital city of Zambia (Unpublished communication with Dr Catherine Chunda-Liyoka). Lusaka is in the central part of Zambia and the SCD numbers are likely to be higher in the northern regions of Zambia.

SCD is therefore, a significant cause of morbidity and mortality in children and young adults in Zimbabwe and Zambia. However, there is paucity of data to allow for accurate characterization of the epidemiology and policy formulation accompanied by limited capacity for diagnosis and clinical management for individuals with SCD. There are also limited national policies or programs to guide surveillance, diagnosis, and management of SCD that have been formulated or implemented. Zambia launched its national clinical guidelines and a pilot newborn screening program for SCD in 2020[17]. In addition, there is a need for an SCD registry and research program to inform future practices and policies in the two countries.

The **S**ickle **H**emoglobinopathy rese**A**rch in **Z**imbabwe and **Z**ambia (SHAZ) program’s overarching goal is to establish a sustainable infrastructure and operation aimed at addressing the paucity of data that currently exists for SCD in the two countries. In addition, the SHAZ program aims to integrate common SCD management guidelines, develop evidence-based care practices, and bring these practices to the bedside via the conduct of implementation science research. Our collaborators include six countries within the SickleInAfrica consortium (Ghana, Mali, Nigeria, Tanzania, Uganda, and South Africa) (https://www.sickleinafrica.org/), the African Institute of Biomedical Sciences and Technology (AiBST) and St Jude’s Children Research Hospital in the USA. The SHAZ program specific objectives are to:

1. Develop and manage an electronic SCD registry with standardized and rigorous clinical follow-up within Zimbabwe and Zambia recruiting at least 4000 participants in 5 years
2. Establish consistent standards of care for patients with SCD in Zimbabwe and Zambia
3. Capacitate investigators to design research on priority areas in SCD relevant to Zimbabwe and Zambia.
4. Establish and improve the biobanking capacity of SCD biospecimens..

We highlight the SHAZ program protocol and lessons learnt in establishing a SCD registry in Zimbabwe and Zambia, two countries within the SickleInAfrica consortium.

## Materials and methodology

### Study design

The SHAZ program involves mixed methods approaches (qualitative and quantitative), that include infrastructure & skilled manpower capacity building of the SCD clinics, a prospective observational registry, biobank, cohort studies to evaluate the effect of various factors in the treatment and management of SCD, and implementation science studies to improve outcomes of SCD. This is a 5-year program extending from 01 May 2021 to 30 April 2026.

### Study setting

Parirenyatwa Teaching Hospital (PTH) in Zimbabwe and UTHs in Zambia are specialized university teaching hospitals that offer a comprehensive care package for children and adults with SCD. Additionally, each hospital has a relatively large bed capacity of 2000.

Zimbabwe: There are 6 implementation sites. The main site is PTH SCD clinic which was the first site to implement the SHAZ program in May 2021. There are 5 outreach sites: Bindura Provincial and Mount Darwin District hospitals in Mashonaland Central Province, Murehwa and Mutoko District hospitals in Mashonaland East Province, and Chinhoyi Provincial Hospital in Mashonaland West Province.

Zambia: There is one site at the UTHs SCD center in Lusaka.

The SHAZ sites have been implementing the registry recruitment, the research projects and standards of care for SCD since project inception. All the sites offer inpatient and outpatient services to patients with SCD.

### SHAZ registry study population

SCD patients from the selected sites in Zimbabwe and Zambia are eligible for recruitment into the SCD registry. The registry recruits all the age groups across the lifespan.

Inclusion criteria

Any participant giving their written informed consent/assent, screened and found to have two abnormal copies of the hemoglobin gene, with at least one of the two genes resulting in production of hemoglobin S:

- Hb SS – sickle cell anaemia
- Hb SC
- Hb Sβ0 thalassemia
- Hb Sβ+ thalassemia
- Hb SD
- Hb SE

We are also including participants with

- Sickle cell trait (Hb AS or another trait variant)

Exclusion criteria:

- Absence of hemoglobin S
- Patients not giving their written informed consent/assent

Only the participants with SCD contribute to the SickleinAfrica consortium registry while the participants with SCT contribute to the local registry data.

Sample size:

Registry aims to recruit at least 4000 participants with SCD in the 5 years of the study.

### Study procedures

Permission was obtained from the Ministries of Health (MOH) to work with PTH, UTHs, Provincial and District health teams to sensitize health workers and raise awareness of the SHAZ program and SCD registry. The Sickle Cell Anaemia Trust in Zimbabwe (SCATZ) and Zambia Sickle Cell Anaemia Society (ZSCAS) are the patient organizations that assist with community engagement activities raising awareness of the SCD registry project and the SHAZ program. Community awareness campaigns have been done using traditional, digital and visual media as well as community events commemorating SCD. Potential study participants are approached in the general pediatric and adult medical wards; the accident and emergency departments; maternity, antenatal and perinatal service units; surgical wards and the outpatient’s clinics (including the vaccination and growth monitoring units).

### Objective 1 and 2

We have set up the SCD center of excellence at PTH and consolidated the one at UTH. We developed the SCD Research Electronic Data Capture (REDCap) database[18] by adopting the Sickle Cell Disease Ontology (SCDO) (https://scdontology.h3abionet.org/) and SIA data elements(https://www.sickleinafrica.org/SIA_data_elements). All SCD participants confirmed SCD by their routine health service providers are approached to seek written consent to participate in the SCD registry. Acceptable laboratory confirmatory diagnostic tests performed before recruitment include sickle cell screen, point of care tests, electrophoresis, isoelectric focusing, High-performance liquid chromatography (HPLC) and / or molecular tests. This is an observational longitudinal registry with no interventions planned which collects information on demographics, clinical information, drug use, treatment and outcomes of children and adults with SCD (see Table 1).

**Table 1.**
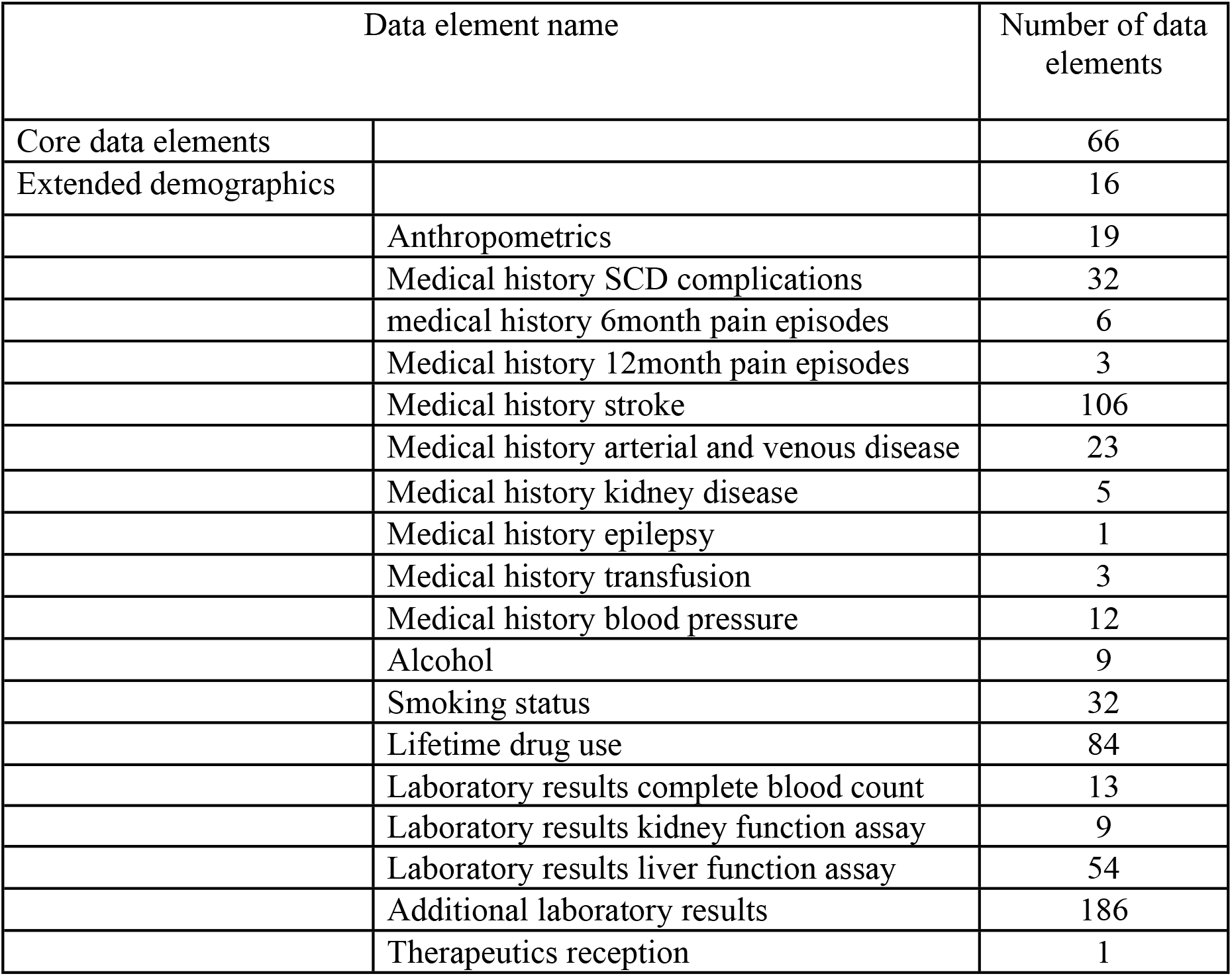
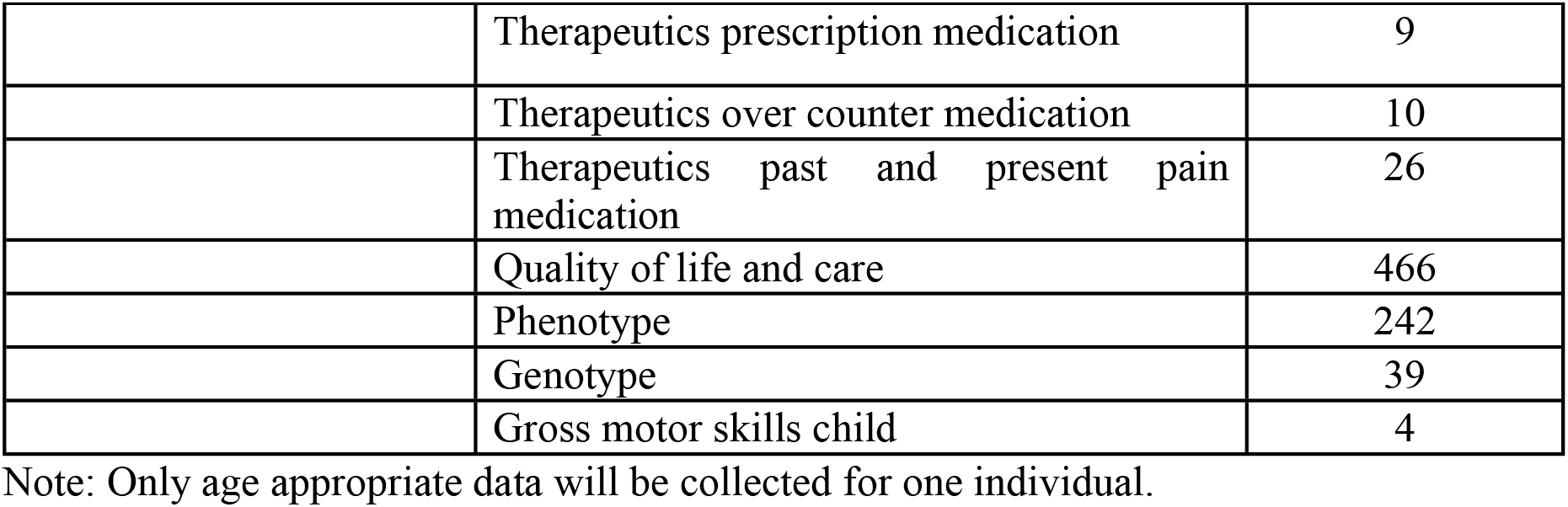
SHAZ registry data elements adopted from the SCDO and SickleinAfrica.

The SHAZ team consisting of the coordinator, outreach workers, and data capture clerks work closely with the recruitment site teams to ensure that participant-related activities and data are promptly entered into the database using mobile devices. Strategic health workers’ training in partnership with the Ministry of Health (MOH) at the two hospitals (PTH & UTH) and outreach sites are continuously being carried out by the SHAZ team to raise awareness of the SCD registry program. All SHAZ recruitment sites are being trained on SCD standards of care adopted from the SicklenInAfrica Consortium[19] (https://sadacc.org/sites/default/files/SPARCo_SoC_SCD_GuidelinesR_Aug2023.pdf), as well as the PEN-Plus program(https://www.ncdipoverty.org/sickle-cell-disease) in collaboration with policymakers in the MOH.

### Objectives 3 and 4

Since the inception of the SHAZ project, we have been working to come up with four consortium-wide studies in the areas prioritized by the SIA consortium countries directly funded by the NIH NHLBI awarded grant. There are two cohort studies on pharmacogenomics of pain management and malaria in children with SCD. There are two implementation studies on newborn screening and penicillin/hydroxyurea use. The four protocols will be implemented by all consortium countries once finalized. We also have been working on various local site studies in both Zambia and Zimbabwe collaborating with MOH, local academic institutions, and other funders. We set up the biorepository for specimen biobanking linked to the proposed cohort and implementation science studies in collaboration with the AiBST in Zimbabwe and the UTH laboratory in Zambia.

### Retention of participants in the SCD registry

The following retention strategies are being applied to retain patients in the registry[20,21]:

### Barrier-reduction retention methods

- Ensuring investigators and support staff are well trained in research methodology, good clinical practice (GCP), and ethics
- Ensuring consistent research staff and hiring staff who can speak the language of the participants as well as understand their cultural values
- Making use of expert SCD patients to mobilize the community and improve community participation.
- Providing SCD education and awareness campaigns and stressing the benefits of participating in long-term cohorts in the communities from which we recruit participants
- Running a pilot test of the registry
- Establishing simple and efficient procedures for data collection. Limiting the number of visits to recruitment and an annual visit for data collection.
- Providing both on-site and telephonic longitudinal data collection to cater to different participants’ preferences. At recruitment, participants are asked to state the method they prefer for communication with the study staff [in person, telephone/cellphone -voice or SMS or social media platforms].
- Ensuring confidentiality and privacy for participants.
- Reimbursing travel costs to participants scheduled for data collection and recognizing the milestones reached in each year of the study.

### Community-building retention strategies

- We have engaged the SCD community advocacy groups (SCATZ & ZSCAS) who provided support letters for the grant application. We will continue to work with them as we implement our program. We have also incorporated them into our Advisory committee which advises the principal investigator and research team.
- Providing education and awareness campaigns on SCD for communities guided by our ministries of health policies and regulations.

### Follow-up and participant reminder strategies

- Using appointment cards and diaries.
- Using telephone or cellular voice or SMS reminders.
- The integration of important or required annual reviews of SCD patients such as urinalysis, BP check, and TCD measurement.

### Tracing strategies

- Documenting alternative contact details of participants.
- Completion of change of address forms at every visit.

## Data management

### Data collection

For the SHAZ registry, data is collected and entered directly into an electronic database developed for the study using REDCap data capture software that is password protected to avoid data compromise. REDCap is an online system that has an embedded mobile application that allows offline collection of data and uploads it later using an internet connection. A data dictionary provided by the SIA Data Coordination Center (DCC) was imported into REDCap. Mobile REDCap data collection using tablet computers was chosen for all recruiting sites and the data capturers have a code name to assist the data manager with resolving data queries. Training on the registry protocol and REDCap software preceded any data collection. All enrolled participants are identified using unique Participant Identifiers (PID) in the database and on data collection forms. All information that identifies the participants such as their names, telephone numbers, and addresses are captured on a separate Study Log kept in a secure place accessed by authorized persons only. Data collectors then upload data through an internet connection to the central server and the Data Manager confirms receipt and reviews the data to check for completeness, validity, and accuracy. If data is collected on paper forms, these forms are reviewed for completeness, accuracy and the use of skip logic before data entry directly into the SHAZ Registry database. Paper forms are stored in secure metal lockable cabinets accessed by the Study Coordinator and Principal Investigator only in both Zimbabwe and Zambia.

All registry data is stored on a password protected local server located in a secure commercial data center in Zimbabwe and data backups are kept at the research sites at PGH and UTH with access limited to the Investigators, Study Coordinator, and Data Manager. Data backup is done on a password protected external media and stored securely at a central place with access limited to only authorized personnel. Data capturers in Zimbabwe have no access to data from Zambia and vice versa. Only the principal investigator, co-principal investigators, study coordinator, and data manager have access to data from both sites. A few data points on gender, age, and date of diagnosis are uploaded to the DCC and this is guided by a data sharing agreement signed by Zambia, Zimbabwe and DCC authorized signatories.

Data quality checks begin during data entry as the database is designed with built-in quality control data entry checks to scan for possible errors, missing data and values out of range. To continuously improve the data quality, integrity and validity, the Data Manager does the Quality Control (QA) real-time review of every single data entry submitted to the central server daily for accuracy, completeness, timeliness, and logic or skip controls. Queries of any data discrepancy noted are sent back to the data collectors for clarification and resolution. The Data Manager produces data quality reports every week and any trends noted in the quality of data are discussed and re-training done to address recurring data errors. Software malfunction issues that arise are discussed with the DCC and the REDCap software developers.

### Data analysis

Summary descriptive statistics will be employed to describe the participants’ socio-demographic, phenotype, and genotype data as well as the prevalence and incidence of complications as we follow up with the participants throughout the study period. The proposed cohort and implementation science studies outlined will have more detailed descriptions once the full protocols are completed at the time of submission for ethics approval for each protocol.

### Ethics

The research regulatory authorities in Zimbabwe, that is, the Joint Research Ethics Committee (JREC/202/21) and Medical Research Council of Zimbabwe (MRCZ/A/2747) provide ethical oversight of this registry and the proposed research activities. The Ministry of Health and Child Care, clinical directors, and respective provincial medical directors approved the research at the PTH and provinces.

In Zambia, ethical approval was sought and granted by the local Research and Ethics committees, of the Excellence Research Ethics & Science Converge, (2021-May-092) and from the National Health Research Authority (NHRA00027/26/09/2023)prior to the commencement of the SHAZ registry program. Permission to conduct the study was also obtained from the Ministry of Health (MOH) and the participating institutional leadership.

SHAZ adopted informed consent forms (ICFs) developed by the CCC to conform with the requirements of the local regulatory authorities. Written informed consent in the participant’s preferred language is obtained using ICFs approved by each country’s ethics committee. A data sharing agreement approved by local ethics committees and institutional leaders was implemented between Zimbabwe and Zambia as well as between SHAZ and the Data Coordination Center. This study is embedded in routine care, there are no physical potential risks rendered to the participants in the registry. However, taking part in the registry may result in people knowing the participant has sickle cell disease and there is a risk the participant may experience social discrimination which can result in psychological distress. Psychosocial support for participants is provided by the SCD clinicians and patient support groups. It is emphasized in the ICF that participation in the registry is voluntary, and participants are free to refuse to participate or withdraw from the registry at any time without loss of benefits to care.

### Timeline of the study

The registry has been recruiting participants as of 1^st^ November 2021 in Zimbabwe and 1^st^ March 2022 in Zambia. A pilot phase was carried out in Zimbabwe from November to December 2021 and any emerging issues were resolved with the DCC. Substantive data collection began in Zimbabwe on 3^rd^ of January 2022 and in Zambia on the 1^st^ of March 2022. Data collection will end on 30^th^ of April 2026 at the end of the funding cycle. The registry project is in the 3^rd^ year of implementation. Table 2 shows the timelines and evaluation metrics for the SHAZ program over 5 years. Year one was a developmental phase of establishing systems for the program linking the two international sites in Zimbabwe and Zambia with the SickleinAfrica consortium already established systems. The registry earnestly began recruitment in the last quarter of year one at both sites and hopefully we will reach 50% or more recruitment by end of year 3.

**Table 2.**
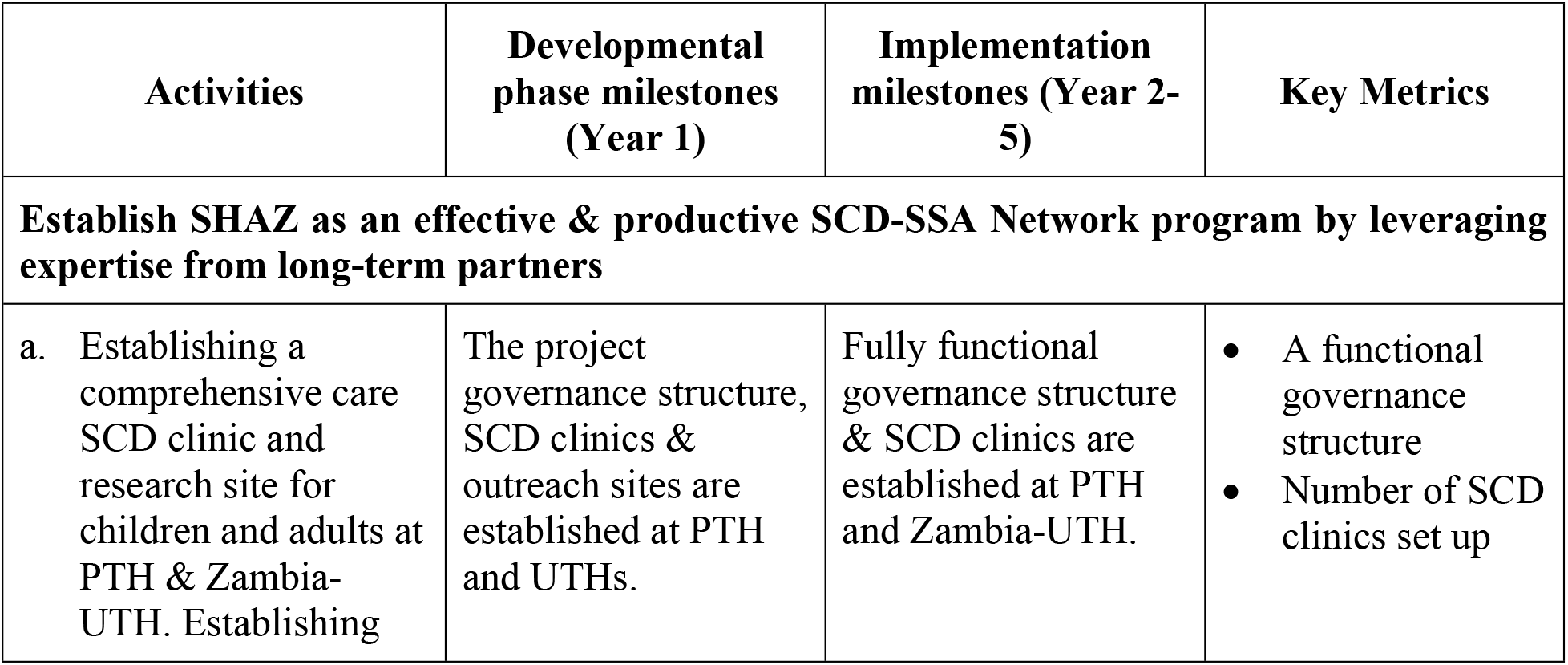
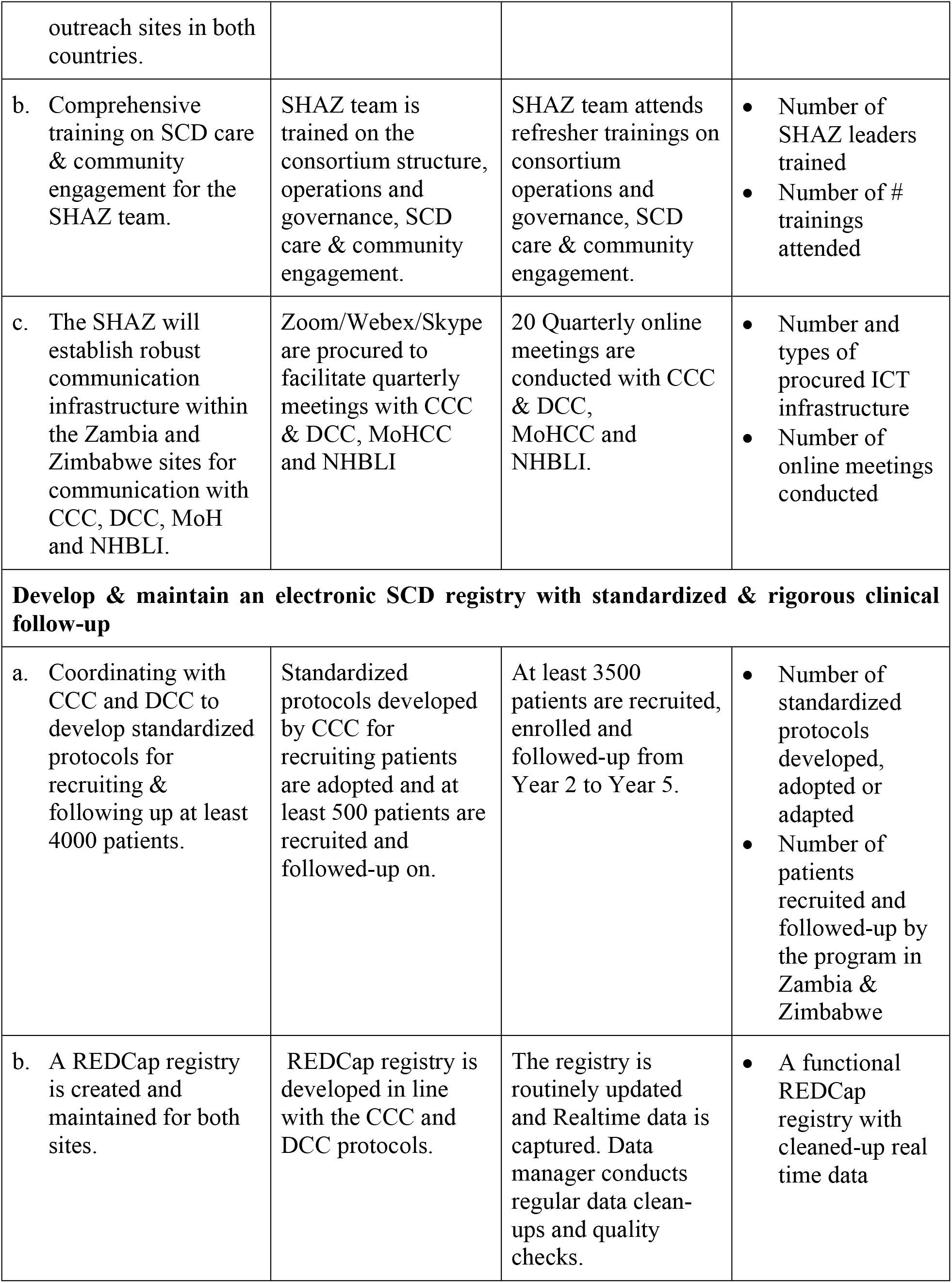
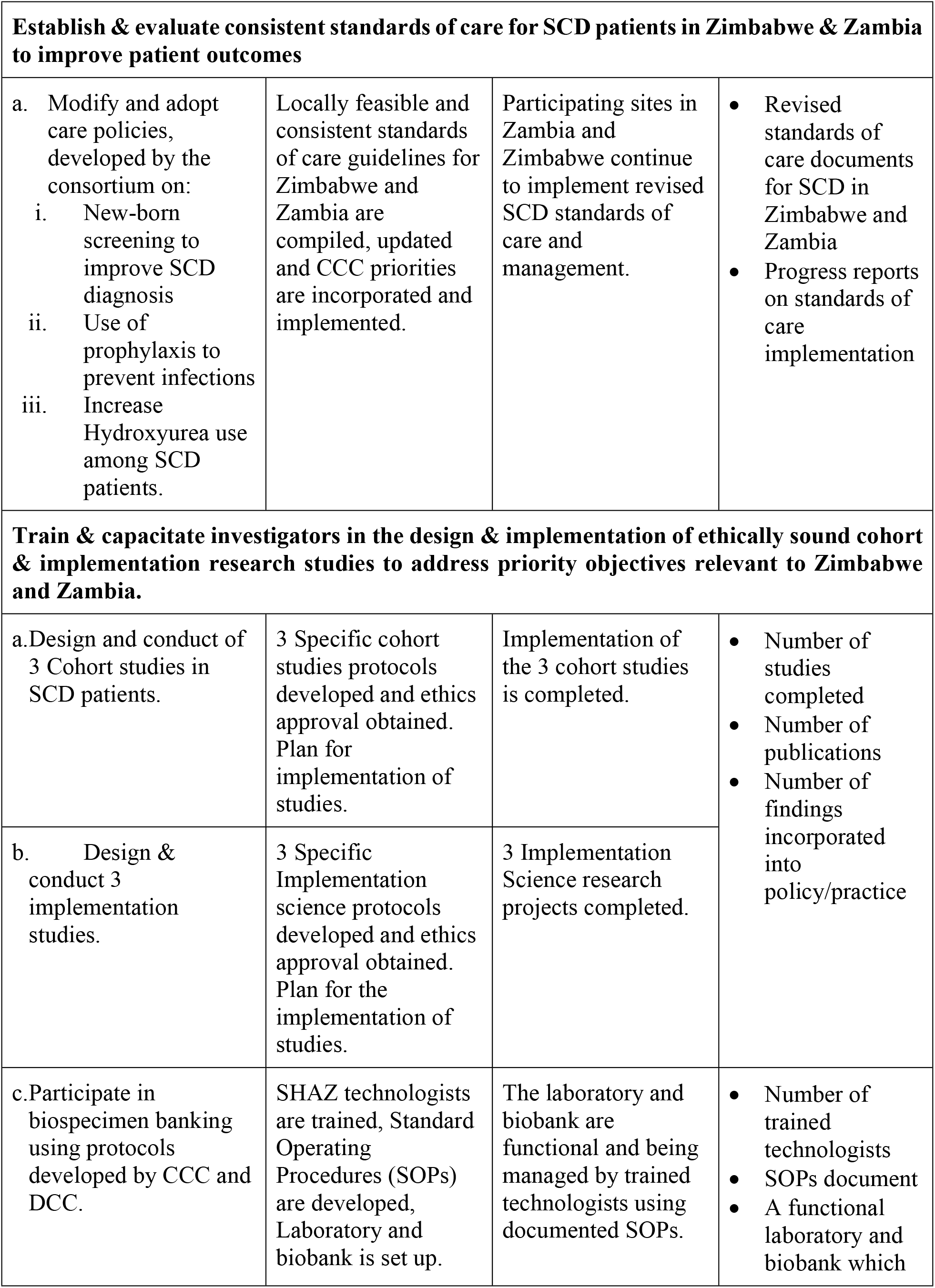
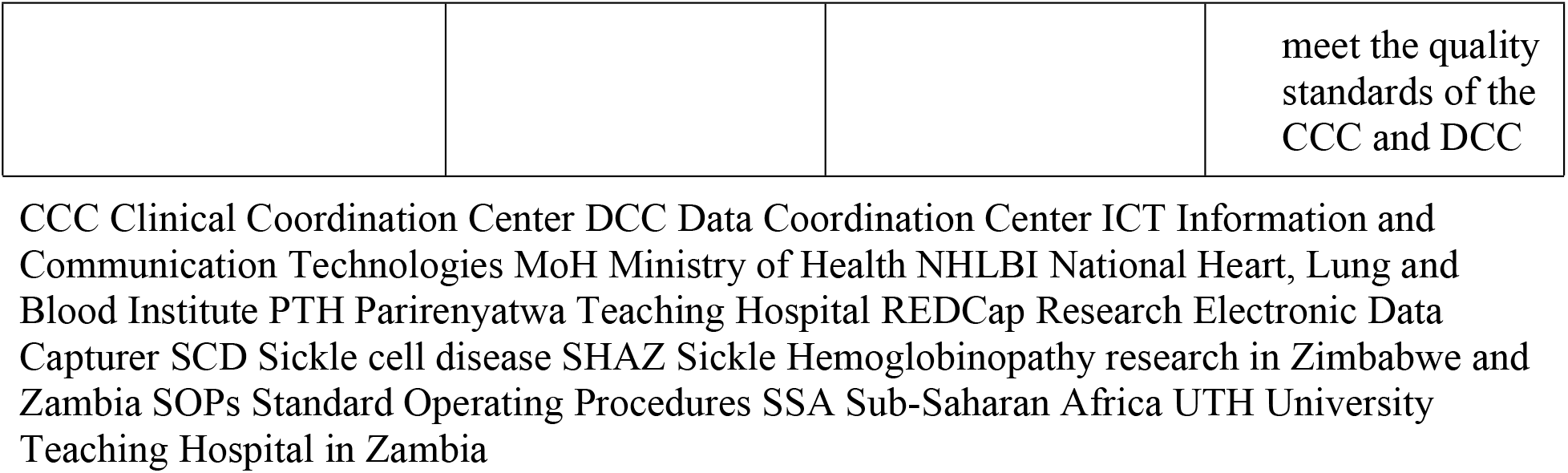
Timelines and evaluation of the SHAZ research activities and outcomes.

| Activities | Developmental phase milestones (Year 1) | Implementation milestones (Year 2-5) | Key Metrics |
| --- | --- | --- | --- |
| **Establish SHAZ as an effective & productive SCD-SSA Network program by leveraging expertise from long-term partners** |  |  |  |
| a. Establishing a comprehensive care SCD clinic and research site for children and adults at PTH & Zambia-UTH. Establishing | The project governance structure, SCD clinics & outreach sites are established at PTH and UTHs. | Fully functional governance structure & SCD clinics are established at PTH and Zambia-UTH. | • A functional governance structure<br>• Number of SCD clinics set up |
| outreach sites in both countries. |  |  |  |
| b. Comprehensive training on SCD care & community engagement for the SHAZ team. | SHAZ team is trained on the consortium structure, operations and governance, SCD care & community engagement. | SHAZ team attends refresher trainings on consortium operations and governance, SCD care & community engagement. | • Number of SHAZ leaders trained<br>• Number of # trainings attended |
| c. The SHAZ will establish robust communication infrastructure within the Zambia and Zimbabwe sites for communication with CCC, DCC, MoH and NHBLI. | Zoom/Webex/Skype are procured to facilitate quarterly meetings with CCC & DCC, MoHCC and NHBLI | 20 Quarterly online meetings are conducted with CCC & DCC, MoHCC and NHBLI. | • Number and types of procured ICT infrastructure<br>• Number of online meetings conducted |
| **Develop & maintain an electronic SCD registry with standardized & rigorous clinical follow-up** |  |  |  |
| a. Coordinating with CCC and DCC to develop standardized protocols for recruiting & following up at least 4000 patients. | Standardized protocols developed by CCC for recruiting patients are adopted and at least 500 patients are recruited and followed-up on. | At least 3500 patients are recruited, enrolled and followed-up from Year 2 to Year 5. | • Number of standardized protocols developed, adopted or adapted<br>• Number of patients recruited and followed-up by the program in Zambia & Zimbabwe |
| b. A REDCap registry is created and maintained for both sites. | REDCap registry is developed in line with the CCC and DCC protocols. | The registry is routinely updated and Realtime data is captured. Data manager conducts regular data clean-ups and quality checks. | • A functional REDCap registry with cleaned-up real time data |

| **Establish & evaluate consistent standards of care for SCD patients in Zimbabwe & Zambia to improve patient outcomes** |  |  |  |
| --- | --- | --- | --- |
| a. Modify and adopt care policies, developed by the consortium on:<br>i. New-born screening to improve SCD diagnosis<br>ii. Use of prophylaxis to prevent infections<br>iii. Increase Hydroxyurea use among SCD patients. | Locally feasible and consistent standards of care guidelines for Zimbabwe and Zambia are compiled, updated and CCC priorities are incorporated and implemented. | Participating sites in Zambia and Zimbabwe continue to implement revised SCD standards of care and management. | • Revised standards of care documents for SCD in Zimbabwe and Zambia<br>• Progress reports on standards of care implementation |
| **Train & capacitate investigators in the design & implementation of ethically sound cohort & implementation research studies to address priority objectives relevant to Zimbabwe and Zambia.** |  |  |  |
| a. Design and conduct of 3 Cohort studies in SCD patients. | 3 Specific cohort studies protocols developed and ethics approval obtained. Plan for implementation of studies. | Implementation of the 3 cohort studies is completed. | • Number of studies completed<br>• Number of publications<br>• Number of findings incorporated into policy/practice |
| b. Design & conduct 3 implementation studies. | 3 Specific Implementation science protocols developed and ethics approval obtained. Plan for the implementation of studies. | 3 Implementation Science research projects completed. |  |
| c. Participate in biospecimen banking using protocols developed by CCC and DCC. | SHAZ technologists are trained, Standard Operating Procedures (SOPs) are developed, Laboratory and biobank is set up. | The laboratory and biobank are functional and being managed by trained technologists using documented SOPs. | • Number of trained technologists<br>• SOPs document<br>• A functional laboratory and biobank which<br>meet the quality standards of the CCC and DCC |

Communication Technologies MoH Ministry of Health NHLBI National Heart, Lung and Blood Institute PTH Parirenyatwa Teaching Hospital REDCap Research Electronic Data Capturer SCD Sickle cell disease SHAZ Sickle Hemoglobinopathy research in Zimbabwe and Zambia SOPs Standard Operating Procedures SSA Sub-Saharan Africa UTH University Teaching Hospital in Zambia

### Plans for dissemination of results

Our target audience includes SCD patient communities, researchers, policy makers, non-governmental organizations and health workers. Table 3 outlines the dissemination plan for the SHAZ Program findings.

**Table 3.** SHAZ program dissemination plan.

| Target group |  | Oral presentations | Written presentations |
| --- | --- | --- | --- |
| SCD patient communities & patient organizations | • ZCAS<br>• SCATZ<br>• Patients in the registry<br>• Other patient organizations in Zimbabwe & Zambia | • Dissemination meetings<br>• Social networks e.g. YouTube, Instagram and X | • Press release<br>• Lay summaries |
| Policy makers | • Ministries of health in Zambia and Zimbabwe | • Dissemination meetings | • Policy briefings<br>• Press release |
| Non-governmental organizations | • Academic institutions | • Dissemination meetings | • Journal articles<br>• Newsletters<br>• Conference posters |
| Health workers | • Public and private institutions in | • Workshops | • Journal articles<br>• Newsletters |
|  | Zimbabwe and Zambia |  |  |
| Researchers | Local, regional and international | Conference oral abstracts | Conference posters<br>Journal articles |

### The current status of the SHAZ Program

1. To establish SHAZ as an effective and productive program in the treatment and management of SCD: The research team engaged and sensitized the policy makers and hospital administrators from both countries on the SHAZ program through an inception meeting and other forums. This has facilitated implementation of the registry, research and standard of care improvement activities in the recruitment sites. Comprehensive care SCD clinics for children and adults with robust outreach programs have been established or improved at the University of Zimbabwe PTH and UTHs in Zambia respectively. SHAZ research team and patient support groups were recruited. The team has received comprehensive protocol training and appropriate training in laboratory skills, genetic counselling, data management, clinical management of patients with SCD and community engagement. We obtained ethics clearance for the SHAZ registry project in both Zimbabwe (JREC/202/21 and MRCZ/A/2747) and Zambia (ERES 2023-Apr-005 and NHRA000006/19/12/2023). We have signed a data sharing agreement between Zimbabwe and Zambia SHAZ programs as well as with the SIA DCC at University of Cape town. We have established 4 out of the 5 outreach sites (Mutoko, Murehwa, Bindura and Mt Darwin) in Zimbabwe and are working on establishing the Chinhoyi site before the end of year 3. We participate in the SHAZ meetings: quarterly Advisory committee meetings, monthly executive committee meetings with Zambia, weekly registry data meetings and weekly site administration meetings. We are also participating in biannually SIA consortium meetings,
2. To develop and maintain an electronic SCD registry with standardized and rigorous clinical follow-up: We adopted the SIA consortium data collection tools to recruit, enroll and follow-up at least 4,000 individuals with SCD. We piloted the registry successfully in year one 3^rd^ quarter and had recruited 48% of participants by February 2024. Zambia has recruited more participants with a ration of 5 to 1 to Zimbabwean numbers reflecting the difference in epidemiology of SCD in the two countries.
3. To establish consistent standards of care for patients with SCD in Zimbabwe and Zambia: We engaged ministries of health technical departments in Zambia and Zimbabwe, evaluated existing standards of care for patients with SCD, adapted SIA consortium and PEN-Plus standards of care recommendations. We have trained 27 health workers from both countries in genomics in 2022. The focus in the 5-year period is on implementing SCD newborn screening, improve use of antibiotic prophylaxis for infection prevention and increased hydroxyurea use among SCD patients.
4. To train and capacity build investigators in the design and implementation of ethically sound cohort and implementation research studies: Our investigators and research staff have participated in training on REDCap software, research methodology for cohort and implementation science research hosted by the CCC, DCC and various partners of the SIA consortium. We have participated in the selection of four SickleInAfrica cross consortium studies:
  - Two cohort studies in the area of SCD pain management pharmacogenomics and Malaria chemoprophylaxis in children with SCD
  - Two implementation studies on SCD newborn screening and Penicillin /Hydroxyurea use SHAZ is leading the development of the SCD pharmacogenomics of pain management study protocol while other consortium members are leading the finalization and implementation of the other studies. The SHAZ lead protocol has been submitted to SIA consortium and NIH for approval. We have also supported site specific research in Zimbabwe (6 students) and Zambia (x students) by masters and doctoral students.
5. To Establish a biobank of the SCD biospecimens : collected and documented in the SCD registry: The SHAZ program has provided additional training to laboratory scientists on biospecimen collection, storage, tracking, shipping and quality control using CCC and DCC protocols. The Biobank has well trained staff made up of 6 people with expertise in management, information technology, ethics, laboratory technologies, and logistics. The biobanking is linked to the research studies. The biobank activities are a collaboration of the SHAZ program, the AiBST and SIA consortium.

## Discussion

The SHAZ program aims to improve SCD SoC through research collaboration with fellow African researchers, policy makers, health workers and SCD patient community groups. It is supporting innovation through research and teaching. We are on track to meet our specific objectives over a 5-year period. The SHAZ registry protocol was amended in Jan 2022 after discussion at SIA consortium level to include an annual follow up for all recruited participants in the registry. Our research strategy was harmonized with the SIA consortium to streamline funding to support four(4) studies (2 cohort and 2 implementation sciences) which will generate more robust evidence on SCD management in Sub-Saharan Africa instead of the originally planned six (6) small studies. These changes though welcome has put a strain on our financial resources and we have expanded our focus to looking for local and international funders to augment our planned research activities. The SHAZ program has created opportunities to network with other SCD African researchers increasing exchange of ideas and expertise. We look forward to the establishment and signing of the SIA charter which will further cement our participation in the consortium activities and sharing of data. We also have benefitted from the CCC and DCC supporting our registry, SCD SoC and research activities through provision of monetary and non-monetary resources. We have recruited two research fellows one in each country.

### Strengths of the SHAZ program

The SHAZ registry is an expansion of the pilot SCD registry that was set up in 2018 at the Parirenyatwa Hospital Paediatric Haematology unit for children under 18 years who have SCD. The registry involves two countries of Zimbabwe and Zambia which share a border on the Zambezi River. Through the SickleInAfrica consortium, Zimbabwe and Zambia have the unique opportunity to learn from their common and distinct experiences to enhance their work on SCD. The establishment of an electronic registry has allowed easy tracking of patients in the registry. A biobank linked to the research studies and registry for both countries was created and this will serve as a reliable resource for future research. The SCD advocacy community groups in both countries are engaged in the process of ensuring support for patients and their families. The Ministries of Health in Zimbabwe and Zambia are supportive of the SHAZ program. The SHAZ program provides technical support for training of healthcare workers to ensure sustainability, improvement of SCD awareness and standards of care. The results from the registry will help to direct policy in SCD diagnosis and management prioritization in the two countries.

Being part of the consortium has facilitated a cascade of training opportunities and activities that may not have been possible for SHAZ to conduct alone. These include SHAZ staff training in data management, genetic counselling, grant writing, research methodology and manuscript writing. The Data Coordinating Center has supported the enrolment of two research fellows in Zambia and Zimbabwe. Collaborating with the SIA consortium is facilitating the development and implementation of consortium-wide studies.

### Limitations of the SHAZ program

The limited diagnostic and management resources for SCD consequently interfere with the identification of affected patients and the provision of appropriate standards of care. There is a need for the research program to find ways of involving other partners to augment funding received and sustain the program. The hospital-based nature of the registry facility excludes patients with asymptomatic or mild disease who do not seek care at the recruitment facilities. However, the involvement of SCD advocacy community groups and the Ministries of Health in both countries helps to increase awareness of the program. This will facilitate recruitment of patients who are not hospital-based. The inconsistency in patient follow-up diminishes the availability of follow-up data for patients and will affect treatment outcome measurement. Only 245 (28 %) of the 853 recruited patients in year one have returned for the one-year follow-up visit. This has highlighted that patients tend to go to the hospital only when they have acute or chronic complications requiring urgent attention. We are working on implementing strategies to increase follow up rates. The cost of hydroxyurea and penicillin V are relatively high for most patients who reside in remote rural areas who are not formally employed. This results in some patients defaulting treatment. There is a need for the program to engage policymakers and other partners to assist in the availability of SCD medications at an affordable price. There is a need for collaborative initiatives with patients and policymakers to lobby for universal treatment for SCD similar to TB and HIV/AIDS programs since SCD is an important non-communicable disease affecting many patients in Sub-Saharan Africa. The study is only covering specific parts of the country, and this limits access for children and adults with SCD in the non-participating regions. However, we hope that engaging the policymakers will have a spillover effect with adoption of policies that will benefit all SCD patients in the two countries.

## Conclusion

The SHAZ program in Zimbabwe and Zambia is already making an impact by engaging policy makers, SCD communities and researchers. At the end of the 5 years, the program will generate hospital based evidence of the burden of SCD in the two countries’ participating sites paving way for bigger and more representative epidemiological research. The SHAZ registry is an important infrastructure which will support basic science, translational and implementation science research. It is raising the awareness for SCD in the two countries and we hope this will translate to improved standards of care for patients with SCD.

## Data Availability

No datasets were generated or analysed during the current study. All relevant data from this study will be made available upon study completion.

## Acknowledgements

We would like to acknowledge the following people:

1. UZ PETRA secretariat: Antony Matsika, Thokozile Mashaah, Nhauro Mupanguri, Felix Madya, Miriro Muvoti, Caroline Tazvivinga and Tendayi Maunganidze (https://petra.org.zw) and Professor James Gita Hakim (posthumously) for the excellent mentorship of the SHAZ research team during the SHAZ protocol and grant writing.
2. The SHAZ advisory committee: Professor Inam Chitsike, Professor Midion Chidzonga, Professor Jonathan Matenga, Dr Nickhill Bhakta, Professor Uma Uthale, Professor Tsungai Chipato, Ms Molyn Chima from SCATZ and Ms Ketty Chunga from ZCAS our SCD community advocates for guiding us through the process of developing and implementing the SHAZ program.
3. The SHAZ research administrator Blessing Murangadzva and Data Capturer Nunurai Sphandla and Driver Mr Jackson Mwepeta for working tirelessly to ensure a smooth implementation of the SHAZ program.
4. The SIA Africa consortium for supporting the SHAZ program participation in the consortium
5. Dr Pauline Kazembe for editing our manuscript language

## References

1. Kanter J, Kruse-Jarres R. Management of sickle cell disease from childhood through adulthood. Blood Reviews. 2013;27: 279–287. doi:10.1016/j.blre.2013.09.001

2. Piel FB, Patil AP, Howes RE, Nyangiri OA, Gething PW, Williams TN, et al. Global distribution of the sickle cell gene and geographical confirmation of the malaria hypothesis. Nature Communications. 2010;1. doi:10.1038/ncomms1104

3. Piel FB, Tatem AJ, Huang Z, Gupta S, Williams TN, Weatherall DJ. Global migration and the changing distribution of sickle haemoglobin: A quantitative study of temporal trends between 1960 and 2000. The Lancet Global Health. 2014;2. doi:10.1016/S2214-109X(13)70150-5

4. Thomson AM, McHugh TA, Oron AP, Teply C, Lonberg N, Vilchis Tella V, et al. Global, regional, and national prevalence and mortality burden of sickle cell disease, 2000–2021: a systematic analysis from the Global Burden of Disease Study 2021. The Lancet Haematology. 2023;10: e585–e599. doi:10.1016/S2352-3026(23)00118-7

5. Wastnedge E, Waters D, Patel S, Morrison K, Goh MY, Adeloye D, et al. The global burden of sickle cell disease in children under five years of age: A systematic review and meta-analysis. Journal of Global Health. 2018;8. doi:10.7189/jogh.08.021103

6. Grosse SD, Odame I, Atrash HK, Amendah DD, Piel FB, Williams TN. Sickle cell disease in Africa: A neglected cause of early childhood mortality. American Journal of Preventive Medicine. 2011;41: S398–S405. doi:10.1016/j.amepre.2011.09.013

7. Wonkam A, Makani J. Sickle cell disease in Africa: an urgent need for longitudinal cohort studies. The Lancet Global Health. 2019;7: e1310–e1311. doi:10.1016/S2214-109X(19)30364-X

8. 60th Session Regional Committee for Africa. SICKLE-CELL DISEASE: A STRATEGY FOR THE WHO AFRICAN REGION. Geneva; 2010 pp. 1–7. Available: https://apps.who.int/iris/handle/10665/1682

9. Piel FB, Rees DC, DeBaun MR, Nnodu O, Ranque B, Thompson AA, et al. Defining global strategies to improve outcomes in sickle cell disease: a Lancet Haematology Commission. The Lancet Haematology. 2023;10. doi:10.1016/S2352-3026(23)00096-0

10. Piel FB, Hay SI, Gupta S, Weatherall DJ, Williams TN. Global Burden of Sickle Cell Anaemia in Children under Five, 2010-2050: Modelling Based on Demographics, Excess Mortality, and Interventions. PLoS Medicine. 2013;10. doi:10.1371/journal.pmed.1001484

11. Piel FB, Patil AP, Howes RE, Nyangiri OA, Gething PW, Dewi M, et al. Global epidemiology of Sickle haemoglobin in neonates: A contemporary geostatistical model-based map and population estimates. The Lancet. 2013;381: 142–151. doi:10.1016/S0140-6736(12)61229-X

12. Pule GD, Chimusa ER, Mnika K, Mhandire K, Kampira E, Dandara C, et al. Beta-globin gene haplotypes and selected Malaria-associated variants among black Southern African populations. Global health, epidemiology and genomics. 2017;2: e17–e17. doi:10.1017/gheg.2017.14

13. Barclay G. Sickle cell anaemia in Zambia. Transactions of the Royal Society of Tropical Medicine and Hygiene. 1971;65: 529–30.

14. Athale UH, Chintu C. The effect of sickle cell anaemia on adolescents and their growth and development: lessons from the sickle cell anaemia clinic. Journal of Tropical Pediatrics. 1994;40: 246–52.

15. Chindima N, Nkhoma P, Sinkala M, Zulu M, Kafita D, Simakando M, et al. The Use of Dried Blood Spots: A Potential Tool for the Introduction of a Neonatal Screening Program for Sickle Cell Anemia in Zambia. Int J Appl Basic Med Res. 2018;8: 30–32.

16. Ministry of Health Zambia. The Zambia Non-Communicable Diseases and Injuries Poverty Commission Report: Reframing Non-Communicable Diseases & Injuries in Zambia. 2022 pp. 19–19. Available: https://static1.squarespace.com/static/55d4de6de4b011a1673a40a6/t/62d0459607789d6c2c9f4b72/1657816491776/Zambia_NCDI+Commission+Report_Final.pdf

17. Makoni M. Newborn screening for sickle cell disease in Africa. The Lancet Haematology. 2021;8: e476–e476. doi:10.1016/S2352-3026(21)00166-6

18. Harris PA, Taylor R, Thielke R, Payne J, Gonzalez N, Conde JG. Research electronic data capture (REDCap)-A metadata-driven methodology and workflow process for providing translational research informatics support. Journal of Biomedical Informatics. 2009;42. doi:10.1016/j.jbi.2008.08.010

19. Paintsil V, Ally M, Isa H, Anie KA, Mgaya J, Nkanyemka M, et al. Development of multi-level standards of care recommendations for sickle cell disease: Experience from SickleInAfrica. Frontiers in Genetics. 2023;13. doi:10.3389/fgene.2022.1052179

20. Teague S, Youssef GJ, Macdonald JA, Sciberras E, Shatte A, Fuller-Tyszkiewicz M, et al. Retention strategies in longitudinal cohort studies: A systematic review and meta-analysis. BMC Medical Research Methodology. 2018;18: 1–22. doi:10.1186/s12874-018-0586-7

21. Robinson KA, Dinglas VD, Sukrithan V, Yalamanchilli R, Mendez-Tellez PA, Dennison-Himmelfarb C, et al. Updated systematic review identifies substantial number of retention strategies: Using more strategies retains more study participants. J Clin Epidemiol. 2015;68: 1481–1487. doi:10.1016/j.jclinepi.2015.04.013.Updated

